# Beyond the Surface: Deep TMS Efficacy in Reducing Craving in Addictive Disorders. A Systematic Review and Meta-analysis

**DOI:** 10.1101/2024.11.13.24317232

**Authors:** Lilia del Mauro, Alessandra Vergallito, Francantonio Devoto, Gaia Locatelli, Gabriel Hassan, Leonor J Romero Lauro

## Abstract

**Background:** Substance use disorders (SUDs) and Gambling Disorder (GD) are addictive disorders with a chronic course. Given the limited efficacy of conventional treatments, there is increasing interest in alternative strategies targeting the altered neural circuits associated with the disease. In this context, deep Transcranial Magnetic Stimulation (dTMS) has emerged as a novel neuromodulation technique capable of reaching deep brain regions. However, no definite recommendation for its use in addiction treatment exists. This study systematically reviewed and quantitatively analyzed dTMS effects in SUDs and GD populations.

**Methods:** Following the PRISMA guidelines, we screened four electronic databases up to February 2024 and selected relevant English-written original research articles. 17 papers were included in the systematic review. As only a minority of studies employed a sham-controlled design, we ran the meta-analysis on a subset of 12 studies, computing the pre-post real stimulation standardized mean change (SMCC) as the effect size, using self-reported craving scores as the dependent variable.

**Results:** The results showed a significant and large effect of active dTMS in reducing craving scores (SMCC = - 1.26, 95% CI [-1.67, - 0.86], *p* <.001). High heterogeneity at both quantitative and qualitative levels across studies was found, with research focusing on different types of SUDs and only one study on gambling behaviors.

**Conclusions:** Results provide initial evidence of the feasibility of dTMS for SUDs care. However, further comprehensive research is needed to unveil several methodological challenges. The limitations of the available literature and future research directions are critically discussed.

## INTRODUCTION

Substance Use Disorders (SUDs) and Gambling Disorder (GD) represent major health concerns worldwide, leading to severe maladaptive consequences at both individual and socioeconomic levels (1,2). SUDs and GD are primary contributors to disability and mortality, substantially increasing the global burden of disease (1,3,4). The prevalence of SUDs and GD is alarming: approximately 284 million people worldwide reported drug use in 2020, reflecting a 26% increase since 2010 (5), whereas 2.3 billion adults engaged in gambling in the past year (6).

The fifth edition of the *Diagnostic and Statistical Manual of Mental Disorders* (DSM-5) classifies both SUDs and GD under the category of substance-related and addictive disorders (7). Similarly, *the International Classification of Diseases*, eleventh edition (ICD-11) (8), categorizes GD as an addictive disorder in light of the growing evidence of behavioral and neurobiological resemblances between the two conditions. At the behavioral level, indeed, both disorders are characterized by a loss of control over the addicted substance or behavior, leading to remarkable distress or impairments in daily life. Addicted individuals typically experience *craving*, an intense urge to consume the substance of abuse or engage in gambling, which has been traditionally linked to relapses and the maintenance of dependence (9–11). This overwhelming desire often leads to neglecting regular activities, severely affecting psychosocial functioning (12). Neurobiologically, addiction is considered a *brain* disorder, characterized by large-scale cortical and subcortical alterations (13,14). Indeed, chronic substance intake induces long-term overactivity in the mesolimbic dopamine system (15), involved in reward and motivational processes, combined with hypoactivity in the frontostriatal circuit (16,17), the neural substrate supporting self-regulation and decision-making functions. Impairment in these processes may account for the weakened ability to stop seeking the substance of abuse (18,19). Importantly, SUDs and GD share partly overlapping structural and functional abnormalities (20,21), particularly in frontostriatal and limbic networks, including the orbitofrontal and anterior cingulate cortex, the insula, the hippocampus, and the amygdala (22).

Although several treatment options are currently available for SUDs and GD, including pharmacotherapy and cognitive behavioral therapy (17,22,23), results remain unsatisfactory, considering retention, dropout, and relapse rates (24,25). Effective alternative or add-on treatment strategies are therefore required. In this context, Transcranial Magnetic Stimulation (TMS) has gained attention as a safe technique for reducing addictive- related symptoms, yielding promising - albeit preliminary - results (26–29).

A significant limitation of standard TMS use in addictive disorders is that figure-of-eight coils can only stimulate surface brain regions, reaching depths of approximately 2 cm beneath the coil. This constraint prevents the possibility of targeting deeper structures critically involved in addictive disorders. To overcome this limitation, Deep TMS (dTMS) has been developed (30). DTMS delivers pulses through the Hedes coil (H-coil), which reaches approximately 4 cm beneath the skull’s surface (31–33) through multiple windings in various planes in the helmet (30), thus enabling stimulation of deep structures such as the thalamus, hippocampus, amygdala, nucleus accumbens, and insula. Specific H-coils have been designed to target different brain networks and, therefore, be applied in specific clinical conditions (for a review, see (31)). For instance, the H1 coil has received FDA clearance for the treatment of major depression, targeting the right and left prefrontal cortex with a preference for the left hemisphere (33). The H4 coil has been approved for smoking cessation, stimulating the prefrontal cortex (PFC) and the insula symmetrically (32). Due to the possibility of targeting bilateral regions and reaching broad and deep subcortical networks, the H-coils have been recently applied in the context of addiction treatment.

Previous systematic reviews and meta-analyses on SUDs and GD analyzed dTMS effects and suggested promising yet preliminary results. However, crucially, those studies typically included only a minority of dTMS studies (34) or explored dTMS effects alongside conventional TMS ones (35,36) or non-invasive brain stimulation (35). To date, only Kedzior et al. (37) systematically analyzed research applying dTMS but evaluated only qualitatively dTMS potential for SUDs care. Therefore, the current study aims to fill the gap in the available literature by providing an updated qualitative synthesis and quantitatively assessing dTMS effects in treating both SUDs and GD populations.

## METHODS AND MATERIALS

### Literature search strategy

Following the Preferred Reporting Items for Systematic Reviews and Meta-Analyses (PRISMA) guidelines (38), we conducted a systematic search on four databases - PubMed, Scopus, EMBASE, and Web of Science - using a combination of terms related to SUDs, GD, and dTMS for articles published up to February 27, 2024. The Supplement reports the search strategy in detail. Papers were included when they: a) were English written, b) involved humans, c) were original research, d) were published in peer-reviewed scientific journals, e) involved patients with SUDs or GD/pathological gambling^1^ diagnosis, f) involved dTMS, and g) dTMS was applied for treatment purposes.

### Record screening

The screening process was run with Rayyan (https://www.rayyan.ai/), a web and mobile systematic review manager (39). After removing duplicates, three blinded authors (G.L., G.H., and F.D.) independently screened the remaining records. The screening included two steps. First, an inclusionary decision was made based on the paper’s title and abstract following the previously described eligibility criteria. Secondly, the same criteria were applied at the full-text level. Conflicts in both steps were resolved by consensus or involving a fourth author (L.D.M). All papers fitting the inclusionary criteria were added to the systematic review and qualitatively synthesized. Studies including sufficient data were also quantitatively analyzed in the meta-analysis. The authors employed two tables, Tables 1 and 2, to extract data from the studies. See the Supplement for details on the literature screening procedure.

**Table 1.** Descriptive information of studies’ samples. AO = add-on; AUD = alcohol use disorder; BD = bipolar disorder; CUD = cocaine use disorder; DD = dysthymic disorder; DSM = Diagnostic and Statistical Manual; dTMS = deep transcranial magnetic stimulation; Hz = hertz; iTBS = intermittent theta burst stimulation; M = male; MD = major depressive episode; MDD = major depressive disorder; NR = not reported; STD = standard detoxification treatment.

| Study | Real dTMS group | Sham dTMS group | Age (mean ± SD) | Education (mean ± SD) | Diagnosis | Medication | Meta-analysis |
| --- | --- | --- | --- | --- | --- | --- | --- |
| Addolorato et al. 2017 (62) | 5 (4 M) | 6 (5 M) | 48.6 ± 9.9 | NR | AUD; DSM-5 Criteria | NR | Included |
| Bolloni et al. 2016 (51) | 10 (9 M) | 8 (7 M) | 33.9 ± 6.5 real group;<br>32.4 ± 10.6 sham group | 11 ± 3.5 real group;<br>11.75 ± 2.31 sham group | CUD; DSM-5 Criteria | NR | Excluded |
| Ceccanti et al. 2015 (63) | 9 (9 M) | 9 (9 M) | 43.22 ± 11.1 real group; 47.29 ± 11.46 sham group | 11.32 ± 3.48 real group; 10.41 ± 3.52 sham group | AUD; DSM-IV Criteria | Not allowed | Included |
| Dinur-Klein et al. 2014 (59) | 7 (4 M) 1 Hz cue group; 7 (3 M) 1 Hz no cue group; 16 (11 M) 10 Hz cue group; 16 (12 M) 10 Hz no cue group | 15 (10 M) cue group; 16 (8 M) no cue group | 48.3 ± 10.8 1 Hz cue group; 50.1 ± 12.1 1 Hz no cue group; 49.9 ± 12.0 10 Hz cue group; 50.3 ± 9.3 10 Hz no cue group; 51.6 ± 10.9 sham cue group; 50.2 ± 7.5 sham no cue group | Primary education (N=4); high-school education (N=46); academic education (N=27) | Tobacco smokers (≥20 cigarettes/day) assessed through phone screening | NR | Included |
| Girardi et al. 2015 (52) | 10 (5M) dTMS-AO group; 10 (7 M) STD group | / | 52.6 ± 7.7 dTMS-AO group; 54.1 ± 11.4 SDT group | ≤ 8 years: 30% dTMS-AO group, 20% SDT group; 9–12 years: 50% dTMS-AO group, 60% SDT group; ≤ 13 years 20% dTMS-AO group, 20% SDT group | DD + AUD; DSM-IV-TR Criteria | Unmodified throughout treatment | Excluded |
| Harel et al.<br>2022 (64) | 23 (15 M) | 23 (15 M) | 43.7 ± 8.7 real group; 42.5 ± 9.8 sham group | 12.5 ± 1.7 real group; 12.2 ± 1.6 sham group | AUD; DSM-5 Criteria | NR | Included |
| Ibrahim et al.<br>2023 (60) | 24 (20 M) | 18 (10 M) | 43.8 ± 12.5 real group, 46.2 ± 12.9 sham group | 14.8 ± 3.6 real group, 14.5 ± 2.8 sham group | Nicotine dependence; DSM-5 Criteria | Patients were provided with varenicline somministratio<br>n | Included |
| Martinez et al.<br>2018 (53) | 6 (6 M) High-frequency group; 6 (6 M) low-frequency group | 6 (5M) sham group | 42 ± 7 high frequency group; 44 ± 5 low frequency group; 44 ± 6 sham group | NR | CUD; DSM-5 Criteria | NR | Excluded |
| Moeller et al.<br>2022 (54) | 10 (7 M) | 10 (7 M) | 50.2 ± 6.8 real group, 47.4 ± 9.9 sham group | NR | Tobacco use disorder + schizophrenia or schizoaffective disorder; DSM-5 Criteria | Unmodified throughout treatment | Excluded |
| Perini et al.<br>2020 (65) | 23 (19 M) | 22 (18 M) | 50.6 ± 10.4 real group, 53.5 ± 7.5 sham group | NR | AUD; Structured Clinical Interview for DSM-IV Axis I (SCID-CV) | NR | Included |
| Rapinesi et al.<br>2013 (67) | 3 (2 M) | / | 52.6 ± 5.5 | 13 | AUD + DD; DSM-IV-TR Criteria and Structured Clinical Interviews for DSM-IV Axis I and II | Unmodified throughout treatment | Included |
| Rapinesi et al.<br>2015 (66) | 12 MDD (7 M); 11 AUD + MDD (6 M) | / | 51.2 ± 8.02 MDD group; 53.6 ± 7.9 MDD + AUD group | 10.7 ± 3.6 MDD group; 10.7 ± 2.6 MDD+AUD group | AUD + MDD Structured Clinical Interviews for DSM- | Stable for at least one month prior to | Included |

**Table 1.** Descriptive information of studies' samples.
|  |  |  |  |  | IV Axis I (SCID-I) and II (SCID-II) | study procedures |  |
| --- | --- | --- | --- | --- | --- | --- | --- |
| Rapinesi et al. 2016 (57) | 7 (7 M) | / | $48.71 \pm 9.45$ | NR | CUD; DSM-IV-TR Criteria | Stable for at least one month prior to study procedures | Included |
| Rapinesi et al. 2018 (55) | 41 (22 M) MDD group; 20 (11 M) BD-I group; 21 (11 M) AUD + MD group | / | $51.44 \pm 10.84$ MDD group; $57.85 \pm 8.19$ BD-I group; $54.38 \pm 7.18$ MD+AUD group | NR | AUD + MD; DSM-5 Criteria | Stable for at least one month prior to study procedures | Excluded |
| Rosenberg et al. 2013 (56) | 5 (5 M) | / | $40.8 \pm 9.4$ | $15.5 \pm 2.5$ | Pathological Gambling; DSM-IV-TR Criteria | One patient was under stable pharmacological medication during treatment | Included |
| Sanna et al. 2019 (58) | 22 (21 M) high-frequency group; 25 (24 M) iTBS group | / | $35.9 \pm 8.5$ high-frequency group; $38.9 \pm 8$ iTBS group | NR | CUD; DSM-5 Criteria | Only some patients were under pharmacological therapy | Included |
| Zangen et al. 2021 (61) | 123 (63 M) | 139 (73 M) | $45.0 \pm 13.0$ real group; $44.8 \pm 13.4$ sham group | Real group: < 9 years = 0%; 9 to 12 years = 33.3%; >12 years = 66.7%. Sham group: < 9 years = 1.4%; 9 to 12 years = 23%; >12 years = 75.5% | Tobacco use disorder; DSM-V Criteria | Psychotropic medications were not allowed | Included |

**Table 2.** Detailed information about dTMS studies protocols. AO = add-on; AUQ = acute urge questionnaire; CCQ-Brief = cocaine craving questionnaire- brief; dACC = dorsal anterior cingulate; DAGS = Dannon and Ainhold gambling scale; DLPFC = dorsolateral prefrontal cortex; dTMS = deep transcranial magnetic stimulation; Hz = Hertz; HF = high frequency; iTBS = intermittent theta burst stimulation; ITI = intertrial interval; LF = low frequency; mPFC = medial prefrontal cortex; NR = not reported; OCDS = Obsessive Compulsive Drinking Scale; PACS = Penn Alcohol Craving Scale ; PFC = prefrontal cortex; rTMS = repetitive transcranial magnetic stimulation; SOGS = South Oaks gambling screen; sTCQ = short form of tobacco craving questionnaire; TCQ = tobacco craving questionnaire; T-QSU = Tiffany questionnaire of smoking urges; VAS = visual analogue scale

| Study | Coil type | Stimulation Site | Stimulation intensity | Stimulation Protocol | Total treatment pulses | Number of sessions | Cue Craving Induction | Craving Measures | Follow-up |
| --- | --- | --- | --- | --- | --- | --- | --- | --- | --- |
| Addolorato et al. 2017 (62) | H-coil (type not specified) | DLPFC, bilaterally | 100% | 3 sessions/week over 4 weeks: 20 trains per session, 50 pulses per train at 10 Hz with an ITI of 15 s | 12000 | 12 | / | OCDS | / |
| Bolloni et al. 2016 (51) | H1 | Bilateral PFC with preference to the left | 100% | 3 sessions/week over 4 weeks: 20 trains per session, 50 pulses per train at 10 Hz with an ITI of 15 s | 12000 | 12 | / | / | 3 and 6 months |
| Ceccanti et al. 2015 (63) | H-coil (type not specified) | Medial PFC | 120% | 5 sessions/week over 2 weeks: 30 trains per session, 50 pulses per train at 20 Hz with an ITI of 30 s | 15000 | 10 | Participants were asked to raise a glass filled with their favorite alcoholic drink, look at it, and sniff it for 5 sec. This action was repeated<br>for 15<br>times in 3<br>min | VAS | Monthly assessments for six months |
| Dinur-Klein et al.<br>2014 (59) | H4 | PFC and<br>insula<br>(bilaterally) | 120% | 5 sessions/week for<br>the first 2 weeks,<br>then 3 non-<br>consecutive<br>treatments on the<br>following week.<br>HF: 33 trains per<br>session, 30 pulses<br>per train at 10 Hz<br>with an ITI of 20<br>sec<br>LF: 600 pulses at 1<br>Hz | 12870<br>(HF);<br>7800<br>(LF) | 13 | Smoking<br>cue | sTCQ | 6 months<br>telephone<br>interview |
| Girardi et<br>al. 2015<br>(52) | H1 | Bilateral<br>PFC with<br>preference to<br>the left | 120% | 5 sessions/week<br>over 4 weeks: 55<br>trains per session,<br>40 pulses per train at<br>20 Hz with an ITI of<br>20 s | 44000 | 20 | / | OCDS | 6 months |
| Harel et al.<br>2022 (64) | H7 | Medial<br>prefrontal<br>and anterior<br>cingulate<br>cortices | 100% | 5 sessions/week<br>over 3 weeks plus 5<br>sessions (1<br>session/week over 5<br>weeks): 100 trains<br>(3 s) per session, 30<br>pulses per train at 10<br>Hz with an ITI of 15<br>s | 60000 | 20 | 3 min of<br>holding<br>and<br>smelling,<br>but not<br>consuming<br>, the<br>alcoholic<br>beverage<br>of choice | PACS |  |
| Ibrahim et al. 2023 (60) | H11 | Insula bilaterally | 120% (110% for individuals with poor tolerability ) | 5 sessions/week over 4 weeks: 34 trains (3 s) per session, 30 pulses per train at 10 Hz with an ITI of 26 s | 20400 | 20 | / | T-QSU | Weekly follow-ups until 8 weeks. Last follow-up at 6 months |
| Martinez et al. 2018 (53) | H7 | mPFC, dACC | 90% on day 1 and then increased to 110% over 2-3 days | 5 sessions/week over 3 weeks. HF: 40 trains per session, 30 pulses per train (3 s) at 10 Hz; 1200 pulses per session LF: 900 pulses at 1 Hz | 15600 (HF); 11700 (LF) | 13 | / | Custom VAS | / |
| Moeller et al. 2022 (54) | H4 | PFC and insula (bilaterally) | 100% on day 1, 110% on day 2, 120% on day 3 | 5 sessions/week over 3 weeks: 60 trains per session, 30 pulses per train at 10 Hz with an ITI of 15 s | 27000 | 15 | / | / | / |
| Perini et al. 2020 (65) | H8 | Insula (bilaterally) | 120% | 5 sessions/week over 3 weeks: 50 trains (3 s) per session, 30 pulses per train at 10 Hz with an ITI of 20 s | 22500 | 15 | 3 min of holding and smelling, but not consuming , a glass of water and the alcoholic beverage choice | AUQ; PACS | 1, 2, 4, 8, and 12 weeks |
| Rapinesi et al. 2013 (67) | H1 | Bilateral PFC with preference to the left | 120% | 5 consecutive sessions/4 weeks: 55 trains (2 s) per session, 40 pulses per train at 20 Hz and with an ITI of 20 s | 44000 | 20 | / | OCDS | 6 months |
| Rapinesi et al. 2015 (66) | H1 | Bilateral PFC with preference to the left | 120% | 5 consecutive sessions/4 weeks: 55 trains (2 sec) per session at 18 Hz and with an ITI of 20 sec | 39600 | 20 | / | OCDS | 6 months |
| Rapinesi et al. 2016 (57) | H1 | Bilateral PFC with preference to the left | 100% | 3 alternate sessions/4 weeks: 20 trains (2 sec) per session, 30 pulses per train at 15 Hz and with an ITI of 20 s, for a total of 720 stimuli per session (total number of trains delivered was 240) | 8640 | 12 | / | VAS | 1 month |
| Rapinesi et al. 2018 (55) | H1 | Bilateral PFC with preference to the left | 120% | 5 consecutive sessions/4 weeks: 55 trains (2 s) per session, 40 pulses per train at 20 Hz and an ITI of 20 s | 44000 | 20 | / | / | 6 months |
| Rosenberg et al. 2013 (56) | H1 | Bilateral PFC with preference to the left | 110% | 1 session/day for 15 days at 1 Hz | NR | 15 | / | SOGS; DAGS; VAS | Families' interviews |
| Sanna et al.<br>2019 (58) | H4 | PFC and<br>insula<br>(bilaterally) | HF: 100%;<br>iTBS:<br>80% | 10 stimulations over<br>the 1st week (2<br>sessions/week-day),<br>4 sessions/ week<br>over the 2nd week,<br>3 alternate sessions/<br>week over the 3rd<br>and 4th week. HF:<br>40 trains per<br>session, 60 pulses<br>per train at 15 Hz<br>with an ITI of 15<br>sec. iTBS: 3 pulses<br>bursts at 50 Hz,<br>repeated at 200 ms<br>intervals for 2 s (i.e.,<br>at 5 Hz). An iTBS<br>train of 2 s was<br>repeated every 10 s<br>for 190 s and 600<br>pulses | 48000<br>(HF);<br>12000<br>(iTBS) | 20 | / | CCQ-Brief | / |
| Zangen et<br>al. 2021<br>(61) | H4 | PFC and<br>insula<br>(bilaterally) | 120% | 5 sessions/week for<br>3 weeks followed by<br>once weekly<br>sessions for 3<br>weeks: 60 trains (3<br>sec) per session, 30<br>pulses per train at 10<br>Hz with an ITI of 15<br>sec | 27000 | 18 | 5 min<br>provocation<br>n<br>procedure:<br>participants<br>were<br>asked to<br>imagine<br>their<br>favorite<br>trigger for<br>craving,<br>viewing images of smoking and listening to audio with instructions to handle a cigarette and a lighter | TCQ;<br>VAS | 3 months |

### Quality assessment

The quality assessment was conducted for all studies included in the systematic review. Four blinded authors (L.D.M., G.L., G.H., and F.D.) assessed the studies’ quality with assessment tools selected based on the study design. Details on the assessment tools, evaluation procedure, and results are reported in the Supplement.

### Quantitative analysis procedure

We extracted relevant information from each article, including dTMS protocol features and sample sizes. Since most of the articles included craving assessment, we selected craving measures as the dependent variable of our analyses. Craving means and standard deviations were collected at baseline, post-treatment, and follow-up. Analyses were run using the “metafor” package for R (version 3.4.3) (40,41). The primary analysis quantified the impact of real dTMS on craving scores, since this stimulation condition was included in all the studies. The standardized mean change (SMCC) (42) was computed as an effect size, including pre and post-treatment craving scores. A secondary analysis was run on the same measure for a subgroup of sham-controlled studies (see Supplement). In this case, Hedge’s g (40,43) was computed as an effect size. For both analyses, we inserted measures so that negative effect sizes indicate a reduction in craving (i.e., an improvement) compared to baseline. Since some studies included more than one effect size, multi-level random effects models were tested (44), and if appropriate, they were reported to handle independence violations (45). Heterogeneity was assessed using several measures, including the Q statistic (for sampling error variation), I² statistic (for variation not due to sampling error) (46), and prediction intervals (PIs) (range where a future observation is likely to fall) (47). We used random-effects models to account for heterogeneity due to sampling error and inter-study variance (48). Subgroup and meta-regression analyses were performed when sufficient data were available (49,50). Publication bias was not analyzed when heterogeneity was high (I² ≈ 75%) (44). For a detailed description of the statistical procedure and analyses of follow-up measures, see the Supplement.

## RESULTS

### Systematic review results Literature search

A total of 2326 records were retrieved from the screened databases. 268 documents were removed as duplicates, and 2058 records were assessed based on their title and abstract following our eligibility criteria. 1961 documents were excluded, while 97 underwent full-text screening. A final sample of 17 papers was considered for the qualitative synthesis. The quantitative analysis was run on 12 studies since 5 (51–55) did not provide sufficient information to be included in the statistical analysis. See Figure 1 for a graphical representation of the screening procedure.

**Figure 1.**
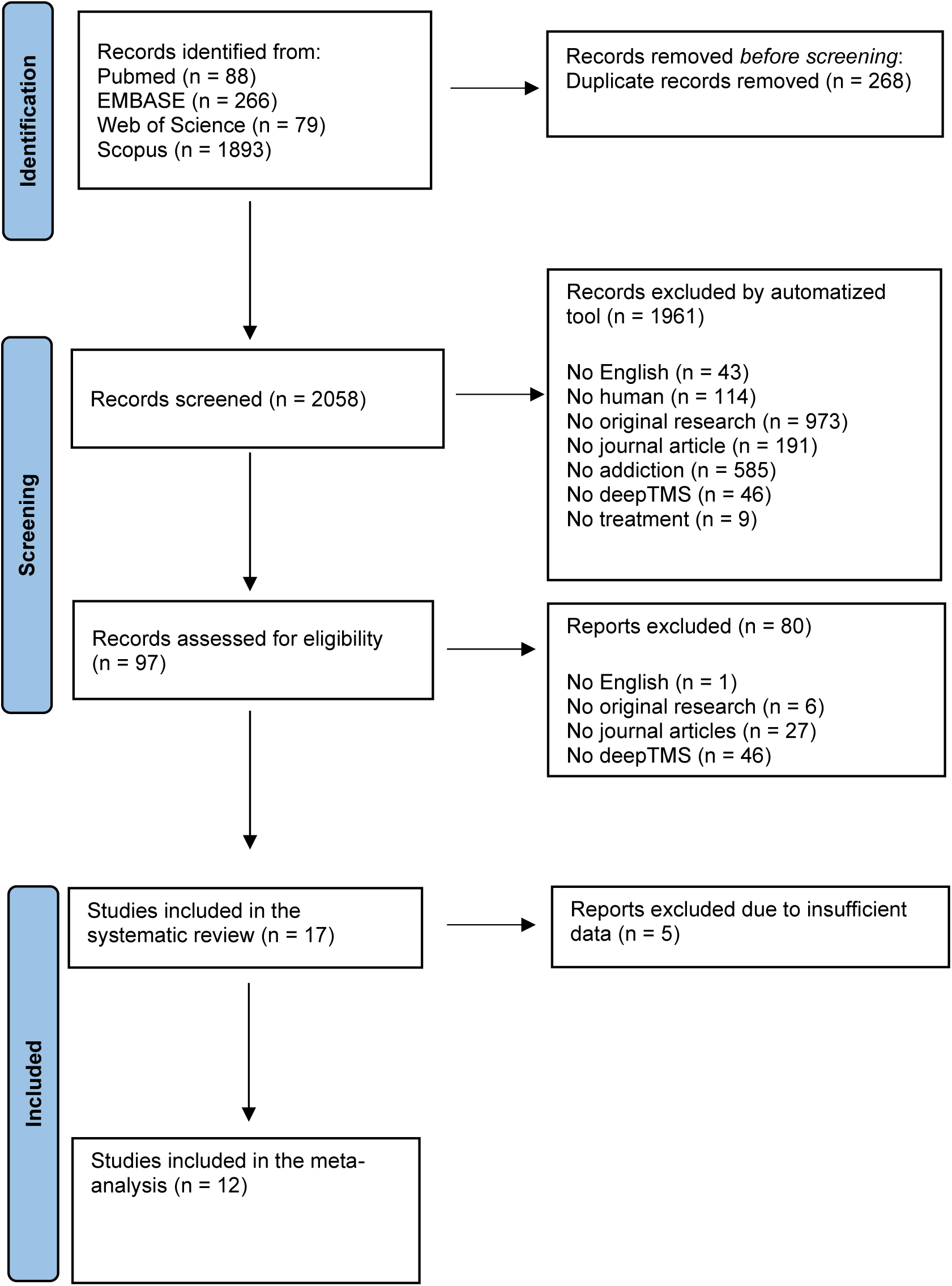
Screening procedure of the selected articles for the systematic review and meta-analysis (38).

### Study details Participants

Our systematic review included 17 studies comprising 747 patients (males = 486, mean age = 46.7 ± 9.3). Substance-related diagnoses included pathological gambling (56), cocaine (51,53,57,58), tobacco (54,59–61), and alcohol (52,55,62–67) use disorders. Diagnoses followed the DSM-IV (63,65,66), DSM-IV-TR (52,56,57,67) or DSM-5 (51,53–55,58,60–62,64) criteria. Psychiatric comorbidities were frequently reported and included dysthymic disorder (52,67), major depressive disorder/episode (55,66), and schizophrenia or schizoaffective disorders (54). In most studies, patients were under a stable pharmacological regimen (52,54–58,60,66,67). See Table 1 for details on the patients’ demographic characteristics.

### DTMS protocols and outcome measures

DTMS protocols were heterogeneous across studies. Most of the research employed the H1 coil, stimulating bilaterally the PFC with a preference for the left hemisphere (51,52,55–57,66,67). Other studies used the H4 coil, targeting bilaterally the PFC and the insula (54,58,59,61). A few studies employed the H8 (65), H7 (53), or H11 (60) coils, stimulating the insula (60,65) or the medial PFC, including the anterior cingulate cortex (53). Moreover, one study (63) employed a specific H-coil version (30) stimulating the medial PFC, whereas one protocol (62) stimulated the dorsolateral prefrontal cortex bilaterally but without specifying the employed coil. 16 groups of patients underwent high-frequency protocols (between 10-20 Hz), whereas low-frequency treatments (i.e., 1 Hz) were employed only in 3 groups of patients (53,56,59). One study (58) included two groups of patients, one undergoing repetitive dTMS and the other dTMS intermittent theta-burst stimulation. The stimulation intensity was set between 80% and 120% of the resting motor threshold. The number of sessions varied from 10 to 20, delivered at different weekly frequencies. 5 studies (59,61,63–65) employed a craving induction procedure before the stimulation, consisting of presenting stimuli aimed at evoking the desire to consume the substance of abuse. The type and modality (e.g., visual, sensory, or imagery) of craving- eliciting procedures varied across studies. Regarding the outcome measures, 14 papers evaluated craving, assessed by either self-report questionnaires or the Visual Analog Scale, whereas one research (55) did not evaluate SUD-related symptoms, Moeller et al. (54) employed a tobacco self-administration task as the outcome measure, and Bolloni et al. (51) used hair analysis to assess treatment efficacy. Concerning objective measures, some studies relied upon measures of substance consumption such as urine (58,59,61,62,64), hair (51), and blood (63) tests. 12 studies included follow-up assessments, which were heterogeneous regarding the time of administration. Studies’ details on treatment protocol and outcome measures are reported in Table 2.

### Pre-post treatment craving scores meta-analysis results

Fourteen effect sizes^2^ were computed. Considering the model selection, the AIC was lower for the reduced model, thus indicating favorable performance, and the LRT comparing the two models was not significant (χ^2^ = 0.70, *p*= .403). Therefore, we selected the reduced one. The results are summarized in the forest plot (Figure 2). The random effects model showed an effect of the treatment in reducing craving scores SMCC = - 1.26, 95% CI [-1.67, - 0.86], significantly different from zero, z = - 6.17, *p* <.001. Therefore, the intervention had a large impact on reducing craving^3^. The meta-analysis also revealed high heterogeneity between studies Q (_13_) = 64.26, *p* < .001, τ^2^ = 0.39 (SE = 0.25) and I^2^ = 79.77% [74.97, 96.98] (substantial heterogeneity), and PIs [-2.70, 0.17]. The Baujat plot inspection (Figure 3) suggested that the effect sizes 8 and 9 (57,66) may be particularly influential considering their impact on estimated heterogeneity and the pooled effect (68). However, the influence analysis did not highlight influential cases. Publication bias was not explored due to the high heterogeneity.

**Figure 2.**
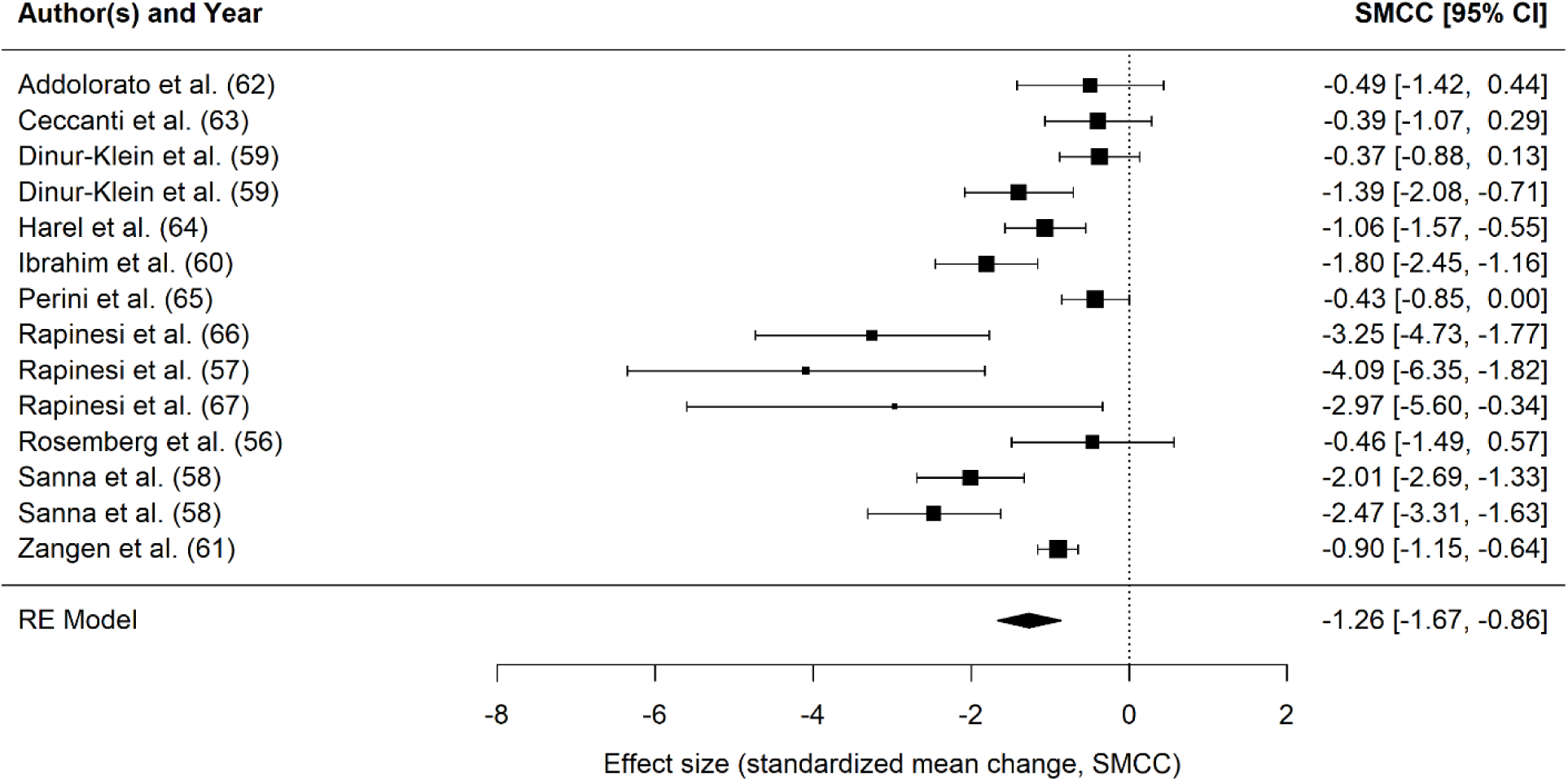
Forest plot summarizing the effect sizes of the dTMS intervention on craving scores. CI = confidence interval.

**Figure 3.**
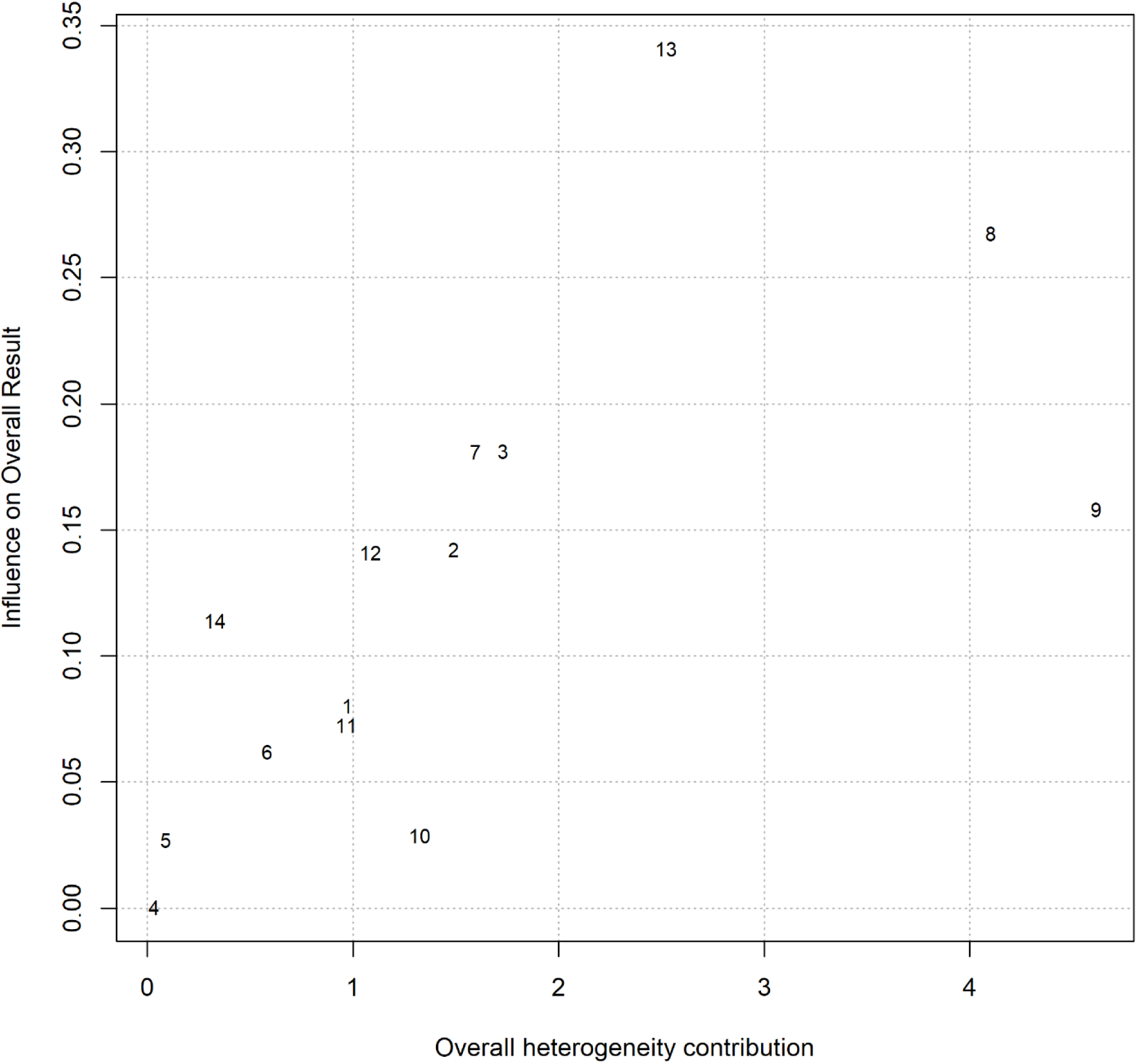
The Baujat plot summarizes the contribution of each study on the meta-analysis heterogeneity considering pre-post craving scores as outcome measure. On the x-axis, the overall heterogeneity as measured by Cochran’s QQ is displayed, whereas on the y-axis, the influence on the pooled effect size is represented.

Considering the meta-regression analysis, we ran a subgroup analysis to explore possible differences in applying H1 vs. H4 coils. Participants’ ages and the number of pulses and sessions were added as continuous predictors. No effects were found for the coil type, patients’ age, or number of pulses (ps > .435). However, the number of sessions significantly predicted SMCC size (p < .001), indicating that craving scores are expected to reduce by increasing the number of sessions. See Supplement for details.

## DISCUSSION

This study systematically reviewed and quantitatively analyzed the effectiveness of dTMS in treating addictive disorders. Seventeen English-written original research articles were included in the qualitative synthesis, and twelve reported sufficient information to run quantitative analyses. All studies but one included SUDs samples; Rosenberg et al. (56) instead included pathological gambling patients. Self-reported craving measures were the most frequent outcome measures across studies. Therefore, they were employed as the primary endpoint of our statistical analyses. Pre to post-real dTMS effect sizes were computed and analyzed in 12 articles, as each included a real stimulation condition. Additionally, since a subset of studies (59–65) included a sham/placebo stimulation, we ran a secondary analysis including only randomized controlled trials (RCTs). The main result of our primary quantitative analysis revealed a significant and large effect of dTMS in reducing craving, aligning with previous findings of standard repetitive TMS (28,34,35,69,70) and dTMS (34–37) in treating addictive disorders. Although previous efforts have already been made to examine dTMS feasibility in addiction care, research in the field shares some limitations. Indeed, Metha et al. (36) did not distinguish dTMS effect sizes from standard rTMS ones, Zhang et al. (34) considered only four dTMS research, Del Mauro et al. (35) analyzed only one dTMS study within non-invasive brain stimulation research, and Kedzior et al. (37) assessed only qualitatively dTMS feasibility in treating SUDs. Therefore, to our knowledge, the current work is the first qualitatively and quantitatively assessing dTMS effectiveness in targeting craving across both SUDs and GD.

Our statistical analysis highlighted a high heterogeneity across studies, which was also observed at the qualitative level. Indeed, studies substantially differed considering participants’ clinical features and stimulation protocols. For instance, SUDs participants varied regarding the preferred substance of abuse, encompassing cocaine, tobacco, and alcohol. Heterogeneity was also observed at the level of comorbid psychiatric disorders and the allowance of concurrent pharmacological treatment. Stimulation protocols differed considering the number of sessions, stimulation frequency, intensity, and type of coil/stimulated regions. Finally, variability was observed considering the specific tool employed to measure craving. We further explored the heterogeneity observed at the qualitative level by running a meta-regression analysis. We did not find differences in effect sizes when using H1 vs. H4 coils. However, considering the restricted number of studies included in the analysis (4 and 5 data points, respectively), caution is warranted in interpreting this finding. Considering the continuous predictors, participants’ age and number of pulses per session did not influence craving scores. Conversely, the increasing number of sessions predicted a larger reduction in craving. The cumulative effect of stimulation sessions aligns with previous studies applying non-invasive brain stimulation across several disorders, including major depressive disorder (71), depressive symptoms in anxiety disorders (72), chronic primary pain (73), and, crucially, substance and eating disorders (69). A crucial point of discussion concerns the variability in brain activity during dTMS administration. Five studies (59,61,63–65) employed a craving induction procedure before dTMS stimulation. The rationale for eliciting craving before stimulation is that brain stimulation effects are known to be state-dependent; that is, the state of the target networks at the moment of stimulation influences brain activity, functional connectivity, and behavioral response (74–77). Therefore, time-locking stimulation with cognitive or behavioral interventions by delivering brain stimulation before, during, or after the stimulation could be beneficial to prime, combine, or consolidate the effects produced by both interventions (78). Moreover, combining brain stimulation with another intervention can be beneficial from another point of view; a key concern in the sole use of brain-based interventions is indeed represented by patients’ expectations of relief. Patients may place responsibility for recovery on the external intervention only, expecting TMS to resolve their addictive behaviors without any personal effort or behavioral changes. In this perspective, multimodal approaches combining stimulation with other interventions avoid this risk, helping patients to keep an active role in abstinence and to develop functional coping strategies. Not surprisingly, a higher internal locus of control is linked to better recovery (79,80). As research in the field advances, dTMS may become an integral part of multimodal treatment strategies for addictive disorders, providing new avenues for managing such complex diseases.

While our results are encouraging, we wish to outline some major challenges in the framework of dTMS administration for addiction care. First, only a minority of studies have been evaluated with a low risk of bias. Therefore, methodological issues such as appropriate sample sizes and blindness must be cautiously considered in future trial designs. Second, most studies employed an open-label design that does not allow disentangling the effects of stimulation from placebo effects. To overcome this limitation in the available literature, we ran a secondary analysis on the subgroup of studies that included a sham-controlled condition. The results revealed a significant small to medium effect of real stimulation in reducing craving, confirming the effectiveness of dTMS. However, the limited number of research included in the analysis suggests caution and highlights the need for more sham-controlled studies to corroborate this finding. As a third point, studies substantially varied regarding the presence of follow-up assessments to track patients’ symptomatology after the end of the treatment, which would provide information considering the durability of induced effects. Despite this variability, we ran an exploratory analysis including follow-up assessments in the real stimulation condition, which suggested a large effect of the intervention on craving reduction compared to the baseline scores. Including follow-up measures can be crucial since, in some cases, rTMS effects (namely the difference of real vs sham stimulation) have emerged only several weeks post-treatment (81–84). This effect might suggest a role of neurostimulation in consolidating long-term neuroplastic change (85).

As a crucial point, craving has been included as the endpoint of our analysis. This choice was data-driven since it was the most reported outcome measure across studies. While craving has been traditionally linked to relapse (10), we acknowledge that other factors, including emotional dysregulation and impulsive decision-making, may drive substance abuse (86–89). Moreover, as a self-report measure, craving is influenced by social desirability and patients’ awareness, varying widely between and within individuals over time and contexts (90). Thus, craving alone may not fully capture consumption behaviors (91), highlighting the need for more objective measures alongside craving assessment. Finally, the inclusion of only one research focusing on gambling signals that research on this disease is still in its infancy, and further exploration is warranted to shed light on dTMS feasibility and effectiveness in treating GD.

### Conclusions

The limited number of included studies, along with sample and protocol heterogeneity and high risk of bias, prevents drawing definitive conclusions on the effectiveness of dTMS in treating addictive disorders. This work represents, however, a first step in understanding the therapeutic impact of dTMS in the context of addiction. Future studies are required to further investigate this desease, providing shared guidelines considering paradigms, outcome measures, and stimulation protocols. Including follow-up measures will also help to clarify dTMS durability after the treatment.

## Data Availability

The study is a meta-analysis, therefore data included and analyzed are already available.

## DISCLOSURES

We declare that L.D.M. works as a psychologist at Fondazione Eris ETS, and L.J.R.L. is a consultant for Fondazione Eris ETS, a non-profit organization dedicated to addiction rehabilitation through a multidisciplinary approach, including the application of dTMS. G.L., F.D., A.V., and G.H., have nothing to declare.

## Footnotes

1 Pathological gambling was the formal diagnosis for GD in the third and fourth editions of the DSM and the tenth edition of the ICD.

2 Twelve studies were included, but two of them (58,59) included two groups each, leading to fourteen effect sizes.

1 The study by Dinur-Klein et al. (59) includes a ‘spurious’ measure of craving. Therefore, we included the study in the meta-analysis but re-run the model without the two effect sizes provided by the study. The statistical results did not change SMCC = - 1.36, 95% CI [-1.82, - 0.90], significantly different from zero, z = - 5.80, *p* <.001.

## Notes

### Funding Statement

This study did not receive any funding

