## Supplementary materials for "Beyond the Surface: Deep TMS Efficacy in Reducing Craving in Addictive Disorders. A Systematic Review and Meta-analysis"

### Supplementary Information

#### Research strategy

Research queries in our databases were built by combining the following keywords: “deep transcranial magnetic stimulation”, “deep repetitive transcranial magnetic stimulation”, “deep rTMS”, “deepTMS”, “deep TMS”, “H-coil”, “substance use disorder”, “SUD”, “addiction”, “gambling disorder”, “cocaine”, “cocaine use disorder”, “CUD”, “tobacco”, “alcohol”, “alcohol use disorder”, “AUD”, “substance dependence”, “substance abuse”, “nicotine”, “drug”, “cannabis”, “smoking”. Table S1 reports the exact search strategy for each screened database and the number of retrieved records.

| **Search databases** | **Search algorithm** | **Records retrieved** |
|---|---|---|
| Pubmed | ("deep transcranial magnetic stimulation" OR "deep repetitive transcranial magnetic stimulation" OR "deep rTMS" OR "deepTMS" OR "deep TMS" OR "H-coil") AND ("substance use disorder" OR SUD OR addiction OR "gambling disorder" OR cocaine OR "cocaine use disorder" OR CUD OR tobacco OR alcohol OR "alcohol use disorder" OR AUD OR "substance dependence" OR "substance abuse" OR nicotine OR drug OR cannabis OR smoking) | 88 |
| Scopus | ALL ( ( "deep transcranial magnetic stimulation" OR "deep repetitive transcranial magnetic stimulation" OR "deep rTMS" OR "deepTMS" OR "deep TMS" OR "H-coil" ) AND ( "substance use disorder" OR sud OR addiction OR "gambling disorder" OR cocaine OR "cocaine use disorder" OR cud OR tobacco OR alcohol OR "alcohol use disorder" OR aud OR "substance dependence" OR "substance abuse" OR nicotine OR drug OR cannabis OR smoking ) ) | 1893 |

| | | |
|---|---|---|
| Embase | ('deep transcranial magnetic stimulation'/exp OR 'deep transcranial magnetic stimulation' OR 'deep repetitive transcranial magnetic stimulation' OR 'deep rtms' OR 'deeptms' OR 'deep tms' OR 'h-coil') AND ('substance use disorder'/exp OR 'substance use disorder' OR sud OR 'addiction'/exp OR addiction OR 'gambling disorder'/exp OR 'gambling disorder' OR 'cocaine'/exp OR cocaine OR 'cocaine use disorder'/exp OR 'cocaine use disorder' OR cud OR 'tobacco'/exp OR tobacco OR 'alcohol'/exp OR alcohol OR 'alcohol use disorder'/exp OR 'alcohol use disorder' OR aud OR 'substance dependence'/exp OR 'substance dependence' OR 'substance abuse'/exp OR 'substance abuse' OR 'nicotine'/exp OR nicotine OR 'drug'/exp OR drug OR 'cannabis'/exp OR cannabis OR 'smoking'/exp OR smoking) | 266 |
| Web of Science | ALL=((“deep transcranial magnetic stimulation” OR “deep repetitive transcranial magnetic stimulation” OR “deep rTMS” OR “deepTMS” OR “deep TMS” OR “H-coil”) AND (“substance use disorder” OR SUD OR addiction OR “gambling disorder” OR cocaine OR “cocaine use disorder” OR CUD OR tobacco OR alcohol OR “alcohol use disorder” OR AUD OR “substance dependence” OR “substance abuse” OR nicotine OR drug OR cannabis OR smoking)) | 79 |

**Table S1.** Details on the search strategy in the screened databases.

### Literature screening procedure

The literature screening procedure entailed labeling each retrieved record according to the selected eligibility criteria as “included,” “excluded,” or “maybe.” The latter label was assigned when a document lacked sufficient information to be excluded or included. The first stage entailed screening papers based on title and abstract only. Documents assessed as “included” or “maybe” were then evaluated based on their full text. In this phase, corresponding authors were contacted in case the articles’ full text was unavailable. During the entire screening procedure, conflicts were solved by consensus or involving a fourth author (L.D.M.).

### Quality of bias tools and assessment procedure

To evaluate the quality of bias of our selected papers, we employed three tools: the Revised Cochrane Risk-of Bias 2 Tool for Randomized Trials (RoB 2) (1) (available at https://sites.google.com/site/riskofbiastool/welcome/rob-

2-0-tool/current-version-of-rob-2?authuser=0), the National Institutes of Health (NIH) quality assessment tool for before–after (Pre–Post) studies with no control group (available at https://www.nhlbi.nih.gov/health-topics/study-quality-assessment-tools), and The Joanna Briggs Institute (JBI) Critical Appraisal Checklist for Case Reports (available at https://jbi.global/critical-appraisal-tools). The former tool evaluates the following bias domains: (a) randomization process, (b) deviations from the intended intervention, (c) missing outcome data, (d) measurement of the outcome, and (e) selection of the reported results. Each domain was rated as "Low," "High," or "Some concerns," and an overall risk of bias judgment of "Low," "High," or "Some concerns" was computed based on the evaluation of the five domains. The NIH tool consists of 12 items evaluating the clarity of the study objective and participants' eligibility/inclusion criteria, the representativeness of the selected sample for the clinical population of interest, the sample size's appropriateness, the description clarity of the delivered intervention, the reliability of the outcome measures employed, the blindness of the people administering the outcome measures for participants' intervention, the potential influence of the loss of assessments after baseline on the results, the appropriateness of the statistical methods, and the presence of follow-ups. Each item was evaluated as "Yes," "No," or "Other". The latter label was assigned in case the specific item could not be determined or applied to the study or when the paper lacked sufficient information to provide a "Yes" or "No" response. Finally, an overall quality judgment equal to "Good", "Fair", or "Poor" was assigned based on the ratings assigned to each item. The JBI Critical Appraisal Checklist for Case Reports is made up of 8 items evaluating the clarity of description of patients' demographic characteristics, history, and current clinical condition, the clarity of diagnostic tests and delivered interventions, the clarity in reporting post-intervention clinical conditions, the clarity in the description of adverse or unanticipated events, and the relevance, in terms of takeaway lessons, of the case report. Each item was evaluated as "Yes," "No," "Unclear", or "Not Applicable". An overall appraisal equal to "Include", "Exclude", or "Seek further info" was assigned based on the ratings assigned to each item. In our quality assessment evaluations, conflicts were solved by consensus.

**Quality assessment results**

Ten studies were randomized controlled trials (2–11) evaluated through the RoB-2. The quality assessment resulted in an overall risk of bias judgment of "some" or "high" concern. Six studies were open-label studies (12–17) evaluated using the NIH. Three papers received a "good," two papers a "fair," and one paper a "poor" overall judgment. Eventually, one research was assessed through the JBI Critical Appraisal Checklist for Case Reports (18), and received an "included" evaluation. Tables S2 – S4 contain details on the quality assessments of the evaluated papers.

| Study | Randomization process | Deviations from Intended Interventions (*effect of assignment to intervention*) | Deviations from Intended Interventions (*effect of adhering to intervention*) | Missing Outcome Data | Outcome Measures | Reported Results Selection | Overall Judgment |
|---|---|---|---|---|---|---|---|
| Addolorato et al., 2017 (2) | Low | Low | Low | Low | Low | Some concerns | Some concerns |
| Bolloni et al., 2016 (3) | Low | Low | Low | Low | Low | Some concerns | Some concerns |
| Ceccanti et al., 2015 (4) | Low | Low | Low | Some concerns | Low | Some concerns | Some concerns |
| Dinur-Klein et al., 2014 (5) | Low | Low | Low | Some concerns | Low | Low | Some concerns |
| Harel et al., 2022 (6) | Low | Low | Low | Low | Low | Some concerns | Some concerns |
| Ibrahim et al., 2023 (7) | Low | Low | Low | Low | Low | Some concerns | Some concerns |
| Martinez et al., 2018 (8) | Low | Low | Low | Low | Low | Some concerns | Some concerns |
| Moeller et al., 2022 (9) | Low | Low | High | Low | Low | Some concerns | High |
| Perini et al., 2020 (10) | Low | Low | Low | Low | Low | Some concerns | Some concerns |
| Zangen et al., 2021 (11) | Low | Low | Low | Low | Low | Some concerns | Some concerns |

**Table S2.** Risk of bias assessment related to the papers evaluated through the RoB-2 tool.

| Study | Overall Judgment |
|---|---|
| Girardi et al., 2015 (12) | Good |
| Rapinesi et al., 2015 (13) | Good |
| Rapinesi et al., 2016 (14) | Fair |
| Rapinesi et al., 2018 (15) | Fair |
| Rosenberg et al., 2013 (16) | Poor |
| Sanna et al., 2019 (17) | Good |

**Table S3.** Risk of bias assessment related to the papers evaluated through the NIH tool.

| Study | Overall Judgment |
|---|---|
| Rapinesi et al., 2013 (18) | Include |

**Table S4.** Risk of bias assessment related to the papers evaluated through the JBI Critical Appraisal Checklist for Case Reports tool.

**Statistical analyses**

We extracted relevant information for each article, comprising dTMS protocol features (coil type, number of sessions, number of pulses), the number of included participants, and their demographic and clinical characteristics. Means and standard deviations were collected from the pre-, post-treatment, and follow-up measurements. In our statistical analyses on follow-ups, measures were included when sufficient data were available, resulting in the analysis of follow-ups performed at 1 (4,7,10,14) and 6 (13,14) months after the end of treatment. Follow-ups were selected to keep homogeneity across studies, considering that follow-ups were performed at different time points. We contacted the authors to obtain the missing data when we found insufficient information in the paper's text, tables, or supplementary materials. When data were available in graphical presentations, we used WebPlotDigitizer (https://plotdigitizer.com/app) to extract them. The primary analysis focused on craving before and after the treatment. Effect sizes were computed as follows: for each included study, we calculated the sampling variance and the standardized mean change (SMCC) using the change score standardization (19), computed with the "escalc" function of the "metafor" package for R (version 3.4.3) (20). We inserted post and pre-treatment values so that negative effect sizes indicate a reduction in craving after the treatment. In contrast, positive values indicate increased craving symptoms after the intervention. The correlation between pre- and post-measurement variances was set at 0.5, as suggested by (21). However, we ran sensitivity analyses establishing lower (0.25) and higher (0.75) correlations to ensure this choice did not influence our results. Considering the included studies, some had sufficient information to calculate more than one effect size per outcome measure (e.g., (5,17)). Considering these effect sizes as statistically independent would violate the independence assumption of traditional meta-analyses and bias the statistical findings (22). Therefore, to address this issue, we ran a multi-level random effects model using the rma.mv function of the "metafor" package, clustering the individual effect sizes at the study level. Then, in line with methodological guidelines (23), we compared the multi-level model with a reduced model (not including the three-level) using the 'anova' function. Based on the Akaike Information Criterion (AIC) and likelihood ratio test (LRT), we selected the best-fitting models and applied them to our data. Fitting three-level models makes sense when it captures data variability more effectively than two-level models. Therefore, we opted for the simpler model when no differences were found between the two (23). The simpler model was run using a random-effects model with the "rma" function. We chose a random-effects model because it is suitable for dealing with heterogeneity due to sampling error and variance between studies' effect sizes (24).

We provided several measures to display data heterogeneity (23). We reported the variation due to the sampling error (Q statistic), the percentage of variation between studies not linked to the sampling error ($I^2$ statistics) (25), and the prediction intervals (PIs) (26), which provide a range into which one can expect the effects of future studies effects to fall based on the available data.

Potential outliers were visually inspected using the Baujat plots (27), a diagnostic tool that allows the detection of studies' contribution to the meta-analyses' heterogeneity. Influential cases were then identified using the influence function 'inf' implemented in the metafor package. As recommended by previous authors (20), when we detected extreme values, we removed them and refitted the model to ensure their elimination did not impact analysis results. Meta-regressions were then run to investigate the effect of moderators and covariates that could explain the heterogeneity and magnitude of extreme values. Potentially interesting moderators and covariates were defined a priori. Moderators included the type of coil (H1, H4) and the specific diagnosis (Alcohol Use Disorder, Cocaine Use Disorder). Covariates comprised the number of sessions, the number of pulses per session, and the participant's age. However, we could not include all the hypothesized factors, but we tested only the informative ones — namely those that were sufficiently represented in the selected papers. As a rule of thumb, Cochrane guidelines suggest that meta-regressions (and subgroup analyses) should be run when at least ten studies are available (28). Considering the limited number of studies in the current work, we performed exploratory meta-regressions when at least four studies per group were available. Considering publication bias analyses, guidelines suggest avoiding them when the between-study heterogeneity is high ($I^2 \approx 75\%$) since results are unreliable (23). When heterogeneity was lower, we explored the publication bias using funnel plots and Egger's regression test (29).

We then included a secondary analysis that considered only randomized controlled trials to investigate whether the effects reported in the previous analysis were due to open-label studies. Differently from the main analysis, we computed the pre-post-treatment mean difference (subtracting pre-treatment measures from post-treatment measures so that a negative value represents symptom improvement) for the experimental and the control group to measure the impact of the non-invasive brain stimulation (NiBS) protocols on the selected outcome measures. We calculated the standard deviation of the pre-post treatment difference according to the Cochrane Handbook for systematic reviews of intervention guidelines (30):

$$SDchange = \sqrt{SDpre^2 + SDpost^2 - (2 * corr * SDpre * SDpost)}$$

Where corr represented the correlation between pre- and post-measurement variances and was set at .5 following (21).

We computed the sampling variance and standardized mean difference (SMD) for each included study using the "escalc" function of the "metafor" package for R, version 3.4.3 (20). The SMD function automatically corrects for the positive bias due to small groups (20,31), computing the Hedge's g (32), used in the present work as an effect size measure. The other steps of the statistical procedure mirrored those previously outlined for the main analysis. The only difference consisted of the Egger regression test: when heterogeneity was lower than $I^2 \approx 75\%$, we used a modified version of the Egger regression test outlined by Pustejovsky and Rodgers (33) (see (23)). Indeed, for SMD effect sizes, the Egger regression test can inflate false positive results due to the non-independence of standardized mean differences and standard errors.

#### Model selection of pre-post treatment craving scores meta-analysis

| Model | df | AIC | LRT | p |
|---|---|---|---|---|
| Full model | 3 | 44.234 | | |
| **Reduced model** | **2** | **42.933** | 0.699 | .403 |

**Table S5**. Summary of the model selection procedure comparing the Full model, namely the three-level regression model, and the reduced model, which does not include the three-level. The best-fitting model is bold.
Note: df = degrees of freedom, AIC = Akaike Information Criterion, LRT = Likelihood Ratio Test.

### Sensitivity analyses

| Correlation | SMCC | SE | CI 95% | Z | p-value |
|---|---|---|---|---|---|
| *Craving pre-post intervention* | | | | | |
| *Corr = .25* | -1.074 | 0.182 | -1.431, -0.718 | -5.909 | <.001 |
| *Corr = .75* | -1.642 | 0.248 | - 2.128, -1.156 | -6.619 | <.001 |
| *Craving pre-follow-up intervention* | | | | | |
| *Corr = .25* | -1.485 | 0.588 | -2.637, -0.333 | -2.526 | .012 |
| *Corr = .75* | -2.096 | 0.827 | -3.717, -0.476 | -2.535 | .011 |

**Table S6.** Sensitivity analysis results. Correlation = values established between pre- and post-measurement variances (0.25, 0.75) (analyses in the main text were run setting a 0.5 correlation); SMCC = standardized mean change (effect size); CI = confidence interval.

### Meta-regression analyses

| Moderator | SMCC | SE | LL | UL | z | p | k | $p_{subgroup}$ |
|---|---|---|---|---|---|---|---|---|
| *Coil type* | | | | | | | | .198 |
| H1 | -2.546 | 0.973 | -4.452 | -0.640 | -2.618 | .009 | 4 | |
| H4 | -1.363 | 0.324 | -1.997 | -0.728 | -4.208 | <.001 | 5 | |

**Table S7.** Categorical moderator and subgroup analysis. Note: SMCC = effect size; SE = standard error of the coefficient; LL = lower limit of the 95% CI; UL = upper limit of the 95% CI; z = z-score associated with the SMCC value in the same row; p = p-value associated with the z-score in the same row; k = the number of effect sizes contributing to SMCC in the same row; $p_{subgroup}$ = p-value of the subgroup comparison.

| Covariate | SMCC | SE | LL | UL | z | p | k | Q | p | $R^2$ (%) |
|---|---|---|---|---|---|---|---|---|---|---|
| *Number of sessions* | | | | | | | 14 | 10.630 | <.001 | 46.64 |
| Intercept | 1.235 | 0.765 | -0.265 | 2.735 | 1.614 | .107 | | | | |
| Slope | -0.153 | 0.047 | -0.246 | -0.061 | -3.260 | <.001 | | | | |
| *Age* | | | | | | | 14 | 0.610 | .435 | 6.26 |
| Intercept | -2.657 | 1.818 | -6.221 | 0.906 | -1.462 | .144 | | | | |
| Slope | 0.031 | 0.040 | -0.047 | 0.109 | 0.781 | .435 | | | | |
| *Number of pulses* | | | | | | | 13 | 0.065 | .798 | 0 |
| Intercept | -1.213 | 0.538 | -2.268 | -0.158 | -2.253 | .024 | | | | |

| | | | | | | |
|---|---|---|---|---|---|---|
| Slope | -0.001 | 0.001 | -0.001 | 0.001 | -0.256 | .798 |

**Table S8.** Summary of the results of continuous moderators on pre-post craving scores. Note: SMCC = Standardized mean change; SE = standard error of the coefficient; LL = lower limit of the 95% CI; UL = upper limit of the 95% CI; z = z-score associated with the SMCC value in the same row; p = p-value associated with the z-score in the same row; k = number of effect sizes contributing to SMCC in the same row; Q = result of the Q-test for moderation; p = p-value of the Q-test for moderation; $R^2$ = amount of heterogeneity accounted for.

In Table S7, the p values in the seventh column show if the subgroup-specific effects are significant. We can see that this is the case for both the studies using H1 and H4 coils. The value under $p_{subgroup}$ shows that the difference in effects between the two subgroups is not significant. These results, however, should be considered exploratory due to the limited number of studies in each group.

Participants' age, the number of sessions, and the number of pulses per session were added as moderators. Results are summarized in Table S8. The meta-regression highlighted that the number of sessions was a significant predictor. For every additional session, the effect size SMCC of a study is expected to decrease by 0.15. Therefore, craving scores in our sample are expected to improve by increasing the number of sessions.

**Craving scores at follow-up**

Six effect sizes were computed measuring follow-up effects. The meta-analysis results are summarized in the forest plot (Figure S1). The random effects model showed an effect of the treatment in reducing craving scores at follow-up compared to the baseline SMCC = -1.71, 95% CI [-3.03, - 0.39], z = - 2.54, p =.011. The meta-analysis also revealed high heterogeneity between studies $Q_{(5)} = 62.56$, $p < .001$, $\tau^2 = 2.26$ (SE = 2.04) and $I^2 = 92.01$ % [74.46, 98.57] (substantial heterogeneity), and PIs [-6.28, 2.86]. The Baujat plot inspection (Figure S2) suggested that the effect size 3 (10) may be particularly influential since it greatly impacts the estimated heterogeneity and the pooled effect. However, the influence analysis did not highlight influential cases. Publication bias was not explored due to the high heterogeneity. Considering the meta-regression analysis, moderators were not included due to the limited number of studies per group. Participants' age, the number of sessions, and the number of pulses per session were added as predictors, but none were significant (see Table S9).

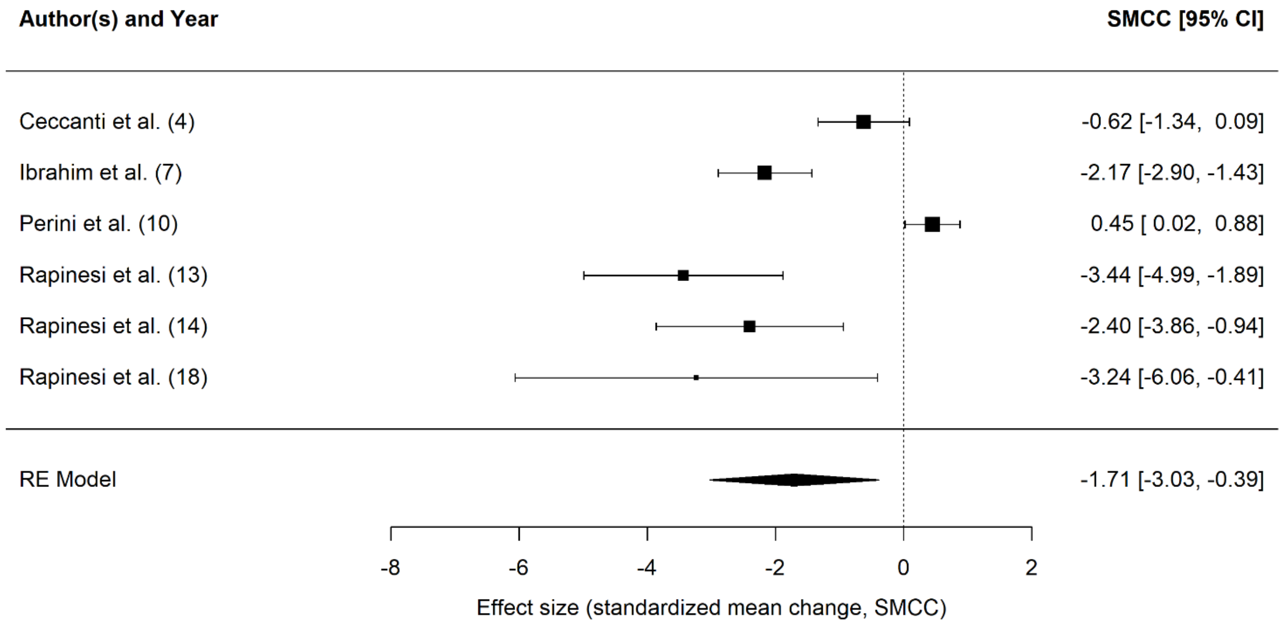

**Figure S1.** The forest plot summarizes the effect sizes of the dTMS intervention on craving scores at follow-up. CI = confidence interval.

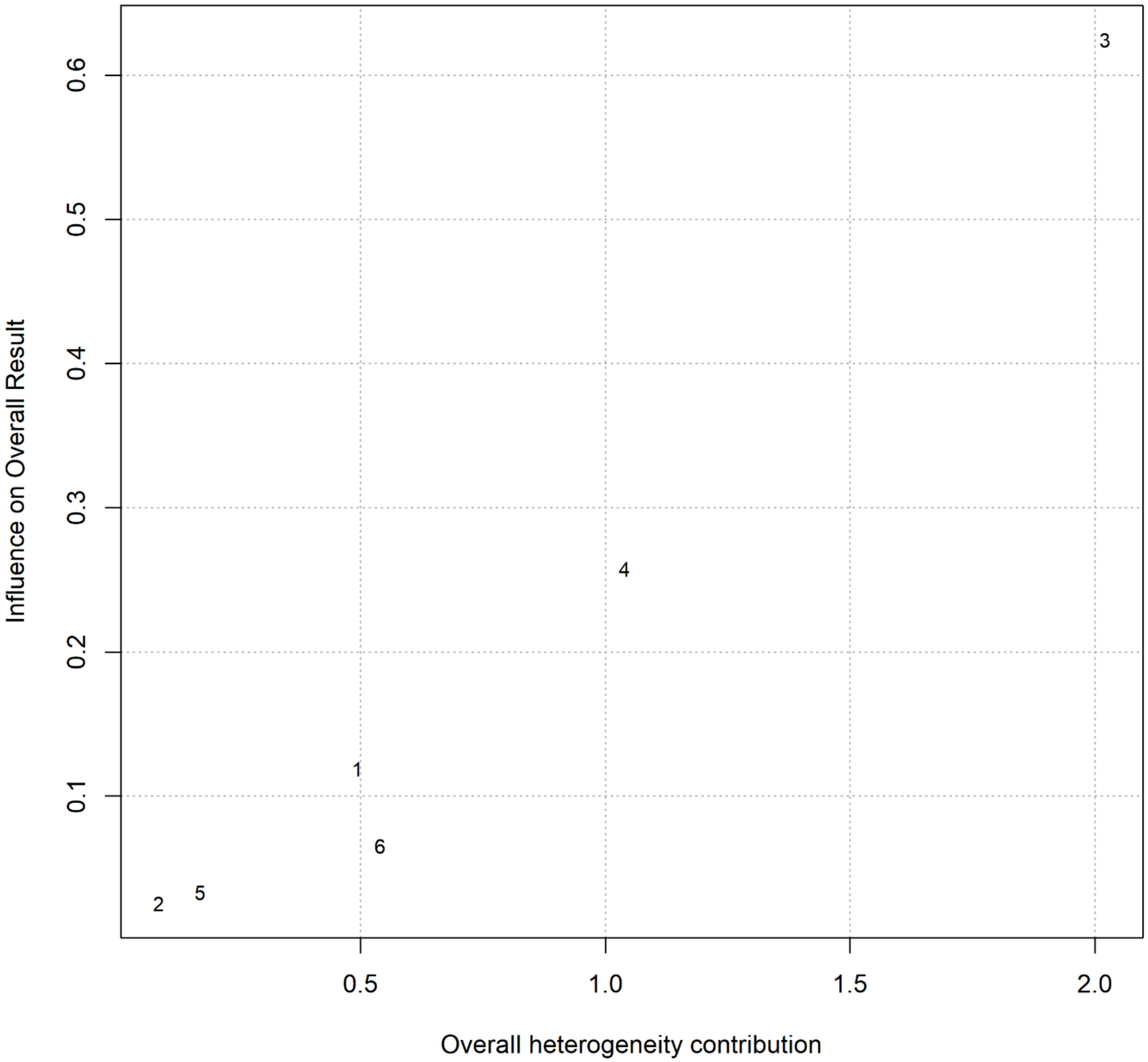

**Figure S2.** The Baujat plot summarizes the contribution of each study on the meta-analysis heterogeneity considering pre to follow-up craving scores as the outcome measure. On the x-axis, the overall heterogeneity as measured by Cochran's QQ is displayed, whereas on the y-axis, the influence on the pooled effect size is represented.

| Covariate | SMCC | SE | LL | UL | z | p | k | Q | p | $R^2$ (%) |
|---|---|---|---|---|---|---|---|---|---|---|
| *Number of sessions* | | | | | | | 6 | 1.349 | .246 | 0 |
| **Intercept** | 1.387 | 2.760 | -4.024 | 6.797 | 0.502 | .615 | | | | |
| **Slope** | -0.196 | 0.169 | -0.527 | 0.135 | -1.161 | .246 | | | | |
| *Age* | | | | | | | 6 | 0.400 | .527 | 0 |
| **Intercept** | 4.193 | 9.435 | -14.298 | 22.685 | 0.444 | .657 | | | | |
| **Slope** | -0.123 | 0.194 | -0.504 | 0.258 | -0.633 | .527 | | | | |
| *Number of pulses* | | | | | | | 6 | 0.126 | .722 | 0 |
| **Intercept** | -.974 | 2.208 | -0.441 | -5.301 | 3.353 | .659 | | | | |
| **Slope** | -0.001 | 0.002 | -0.003 | 0.002 | -0.356 | .722 | | | | |

**Table S9**. Summary of the results of continuous moderators on pre to follow-up craving scores. Note: SMCC = Standardized mean change; SE = standard error of the coefficient; LL = lower limit of the 95% CI; UL = upper limit of the 95% CI; z = z-score associated with the SMCC value in the same row; p = p-value associated with the z-score in the same row; k = number of effect sizes contributing to SMCC in the same row; Q = result of the Q-test for moderation; p = p-value of the Q-test for moderation; $R^2$ = amount of heterogeneity accounted for.

#### Pre-post treatment craving scores in sham-controlled studies results

Eight effect sizes[1] were included in the analysis. The model comparison indicated a lower AIC for the reduced model, thus indicating favorable performance, whereas the LRT comparing the two models was not significant ($\chi^2_{(1)} = 0$, p=1). Therefore, we selected the reduced model. The results are summarized in the forest plot (Figure 4). The random effects model showed an effect of the treatment in reducing craving scores g = - 0.35, 95% CI [-0.54, - 0.15], significantly different from zero, z = - 3.47, p < .001. Hence, the real stimulation had a large impact on reducing craving[2]. The meta-analysis did not reveal heterogeneity between studies $Q_{(7)} = 6.80$, p = .450, $\tau^2 = 0$ (SE = 0.05), $I^2 = 0\%$ [0, 79.47] (no heterogeneity), and PIs [-0.59, -0.10]. The Baujat plot inspection (Figure 5) suggested that the effect sizes 6 (7) and 3 (5). The influence analysis, however, highlights the effect size 8 (11) as an influential case, probably due to its larger sample size than the other studies (83 participants in the real condition vs. 106 in the sham). However, the removal of this study did not impact the results g = -0.30 95% CI [-0.58, -0.02], which is significantly different from zero, z = - 2.10, p =.036. Therefore, the effect of real stimulation remained significant even when including the sham/placebo condition in the analyses.

Considering the publication bias, the modified Egger test showed no asymmetry b = -0.66, 95% CI [-2.56, 1.23], z = 0.35, p = .723.

[1] Seven studies were included, but (5) included two different groups, leading to eight effect sizes.

[2] The study by Dinur-Klein et al. (5) includes a 'spurious' measure of craving. Therefore, we included the study in the meta-analysis but re-run the model without the two effect sizes provided by the study. The statistical results did not change SMCC = - 0.32, 95% CI [-0.53, - 0.10], significantly different from zero, z = - 2.92, p =.004.

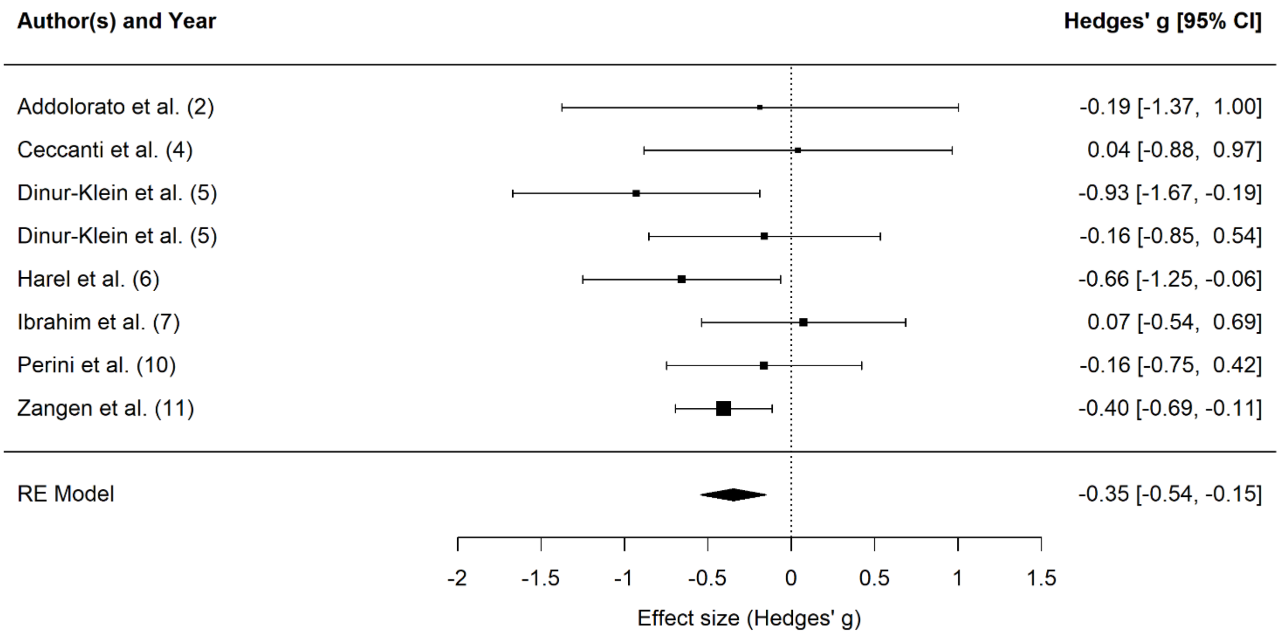

**Figure 4.** The forest plot summarizes the effect sizes of the dTMS on craving scores in a subgroup of studies including a sham condition. CI = confidence interval.

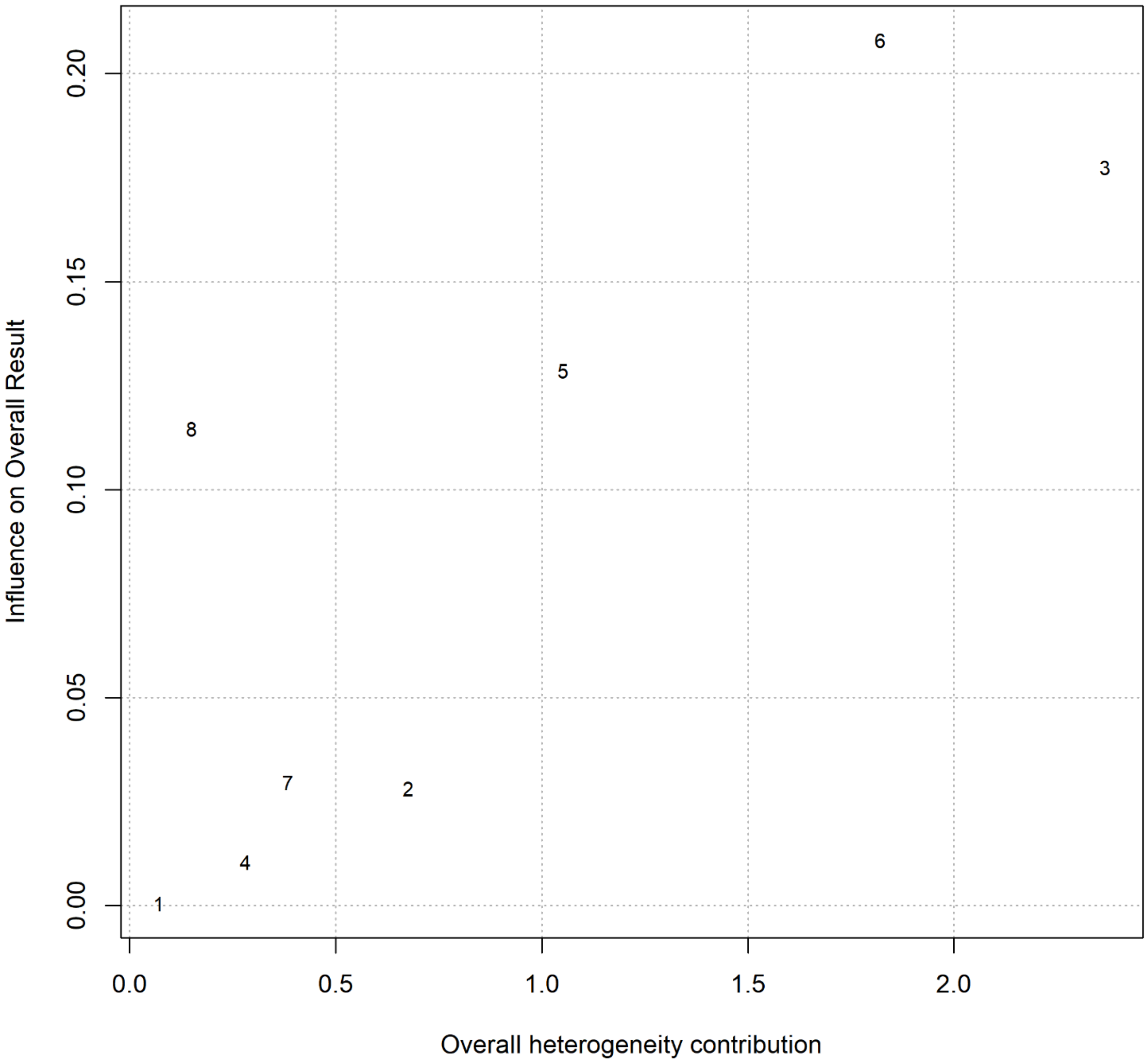

**Figure 5.** The Baujat plot summarizes the contribution of each study on the meta-analysis heterogeneity, considering pre- to post-craving scores as the outcome measure in a subgroup of studies including a sham/control condition. On the x-axis, the overall heterogeneity as measured by Cochran's QQ is displayed, whereas on the y-axis, the influence on the pooled effect size is represented.
